# Inference of naturally-acquired immunity using a self-matched negative control design

**DOI:** 10.1101/2020.03.01.20029850

**Authors:** Graham R. Northrup, Lei Qian, Katia Bruxvoort, Florian M. Marx, Lilith K. Whittles, Joseph A. Lewnard

## Abstract

Host adaptive immune responses may protect against infection or disease when a pathogen is repeatedly encountered. The hazard ratio of infection or disease, given previous infection, is typically sought to estimate the strength of protective immunity. However, variation in individual exposure or susceptibility to infection may introduce frailty bias, whereby a tendency for infections to recur among individuals with greater risk confounds the causal association between previous infection and susceptibility. We introduce a self-matched “case-only” inference method to control for unmeasured individual heterogeneity, making use of negative-control endpoints not attributable to the pathogen of interest. To control for confounding, this method compares event times for endpoints due to the pathogen of interest and negative-control endpoints during counterfactual risk periods, defined according to individuals’ infection history. We derive a standard Mantel-Haenszel (matched) odds ratio conveying the effect of prior infection on time to recurrence. We compare performance of this approach to several proportional hazards modeling frameworks, and estimate statistical power of the proposed strategy under various conditions. In an example application, we use the proposed method to re-estimate naturally-acquired protection against rotavirus gastroenteritis using data from previously-published cohort studies. This self-matched negative-control design may present a flexible alternative to existing approaches for analyzing naturally-acquired immunity, as well as other exposures affecting the distribution of recurrent event times.

## INTRODUCTION

Host adaptive immune responses often protect against infection or disease when a pathogen is repeatedly encountered. Vaccines aim to exploit this mechanism of protection by exposing hosts to an attenuated infection, or to immunizing subunits of a pathogen. As such, evidence of protective naturally-acquired immunity provides strong rationale for vaccine development.^1^ Quantitative estimates of the strength of naturally-acquired protection also inform the interpretation of epidemiologic data, for instance providing a baseline against which vaccine performance can be evaluated.^2^ These estimates are further sought to parameterize mathematical models of pathogen transmission.^3^

Naturally-acquired immunity is often estimated via the hazard ratio of infection or disease, comparing counterfactual periods representing person-time at risk in the presence and absence of prior infection.^4–10^ Thus, inference centers on the distribution of recurrent event times. Unmeasured heterogeneity in individuals’ hazard rates of infection or disease presents a challenge in such analyses, originally termed a problem of “varying liabilities” by Greenwood and Yule^11^ and subsequently addressed as “accident-proneness”^12^ or “frailty”.^13^ The tendency for events to recur among certain individuals must be accounted for in statistical analyses.^14^ For instance, in studies of naturally-acquired immunity, recurrence of infection or disease among individuals with the greatest susceptibility or exposure to a pathogen, irrespective of previous infection, may bias estimates of naturally-acquired protection.^15^

This consideration may have relevance to several diseases against which immune responses are thought to generate imperfect protection. Tuberculosis presents a notable example, where despite evidence of protective cell-mediated and humoral immunity,^16^ several epidemiologic studies have reported higher rates of new-onset infection or disease among persons previously treated successfully for active tuberculosis, as compared to those without history of tuberculosis.^17–20^ Similar conflict about the consequences of prior infection has arisen in epidemiologic studies of gonorrhea.^21,22^ In recent analyses of a multi-site pediatric cohort study addressing enteric disease, previous infection predicted higher rates of recurrent infection or disease associated with several pathogens, including *Shigella* spp., *Campylobacter* spp., and various diarrheagenic *Escherichia coli* strains.^23^ Evidence supporting the feasibility of protective vaccines against many of these pathogens suggests a need to revisit the impacts of naturally-acquired immunity.^24–26^ Similar causal inference challenges arise in the relationship between chronic inflammation and repeated infection in conditions such as cystic fibrosis,^27,28^ otitis media,^29,30^ and environmental enteric dysfunction.^31^

Formalizing unmeasured heterogeneity as a problem of confounding suggests potential strategies to identify naturally-acquired protection. Terming *Y*_1_ and *Y*_2_ as primary and recurrent infection or disease outcomes, respectively, and *U* as the constellation of unmeasured individual factors influencing exposure or susceptibility to a pathogen of interest, a directed acyclic graph (**Figure 1**) reveals that *Y*_1_←*U*→*Y*_2_ may introduce bias into estimation of the causal relationship of interest, *Y*_1_→*Y*_2_. Conditioning on unmeasured individual factors by comparing observations during counterfactual risk periods from the same individual (*Y*_1_←*U*→*Y*_2_) permits unbiased inference of the effect of *Y*_1_. This intuition provides the basis for numerous self-matched designs (e.g. case-crossover, case-time control, and self-controlled case series), which have garnered increasing interest in epidemiology in recent years.^32^

**Figure 1:**
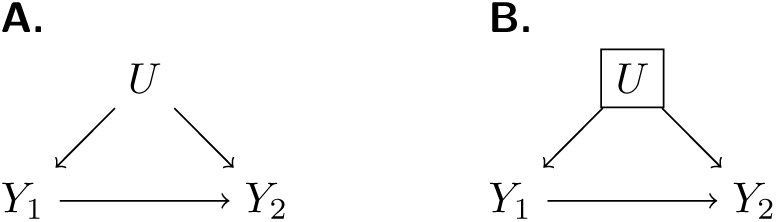
Directed acyclic graph addressing unmeasured confounding. We illustrate a causal framework wherein the effect of previous infection on time to subsequent infection (*Y*_1_→*Y*_2_) is of interest for analysis. One or more unmeasured confounding factors (*U*) creates a backdoor path (**A**) which can be blocked by conditioning on *U* (**B**).

In this paper, we present an adaptation of these methods harnessing data from “negative control” events to permit causal inference in the presence of heterogeneous individual frailty. We derive a matched (Mantel-Haenszel) odds ratio (*OR*_*MH*_)^33,34^ estimator for the hazard ratio of infection or disease, given previous infection. We conduct simulations to compare this approach against alternative methods based on proportional hazards models common in the analysis of longitudinal data, and to assess statistical power under varying conditions. Last, we use the proposed method to reassess protective effects of rotavirus infection in data from previously-published birth-cohort studies.^4,5^

## APPROACH

### Self-matched negative control design

Consider an outcome such as acquisition of a pathogen of interest, or onset of disease due to this pathogen (**Table 1**). The proposed design only includes individuals who experience recurrent episodes of this outcome of interest (case-only). Define *Y*_*i*_ and *X*_*i*_ as variables indicating outcome and exposure status for individual *i* at each observation, with *Y*_*i*_ = 1 indicating infection or disease with the pathogen of interest and *Y*_*i*_ = 0 indicating a negative-control outcome. Consideration of negative-control observations is of interest for studies involving event-based data capture (e.g. episodes of acute illness), and provides a basis for a competing risks estimation framework as we detail below. Last, let *X*_*i*_ = 1 indicate an individual has previously experienced infection with the pathogen of interest, and with *X*_*i*_ = 0 indicating the individual has no history of infection with the pathogen of interest.

**Table 1:** Parameters and definitions.

| Parameter | Definition |
| --- | --- |
| $\lambda_{Pi}$ | Rate at which individual $i$ experiences a pre-specified clinical endpoint due to the pathogen of interest (“outcome of interest”), in the absence of naturally-acquired immunity |
| $\lambda_{Ni}$ | Rate at which individual $i$ experiences a negative-control outcome |
| $\theta$ | Hazard ratio for the outcome of interest, owing to naturally-acquired protection |
| $\beta_{Pi}(1)/\beta_{Pi}(0)$ | Hazard ratio for the outcome of interest during the period after primary infection, relative to the period before primary infection, for individual $i$, due to all (confounding) factors other than naturally-acquired protection |
| $\beta_{Ni}(1)/\beta_{Ni}(0)$ | Hazard ratio for the negative control outcome during the period after primary infection, relative to the period before primary infection, for individual $i$ |

Define *A*_*i*_-*D*_*i*_ as random variables indicating event times for observations of *Y*_*i*_ = 1 and *Y*_*i*_ = 0, conditioned on *X*_*i*_, according to the contingency structure presented in **Table 2**. *A*_*i*_ and *B*_*i*_ are the time to first occurrence of the outcome of interest and the negative control outcome, respectively, for an individual with no history of infection (*X*_*i*_ = 0). *C*_*i*_ and *D*_*i*_ are the time to the first occurrence of the outcome of interest and the negative control outcome, respectively, following infection with the pathogen of interest (such that *X*_*i*_ = 1; **Figure 2**). Here we note that *B*_*i*_ and *D*_*i*_ are censored if *A*_*i*_ < *B*_*i*_ and *C*_*i*_ < *D_i_*, respectively.

**Table 2:** Contingency table for event time distributions, given prior infection.

| Exposure status |  | Outcome status |  |
| --- | --- | --- | --- |
| | | Outcome of interest<br>$Y_i = 1$ | Negative control outcome<br>$Y_i = 0$ |
| Previously uninfected | $X_i = 0$ | $A_i \sim \text{Exp}(\lambda_{Pi})$ | $B_i \sim \text{Exp}(\lambda_{Ni})$ |
| Previously infected | $X_i = 1$ | $C_i \sim \text{Exp}(\theta \lambda_{Pi})$ | $D_i \sim \text{Exp}(\lambda_{Ni})$ |

**Figure 2:**
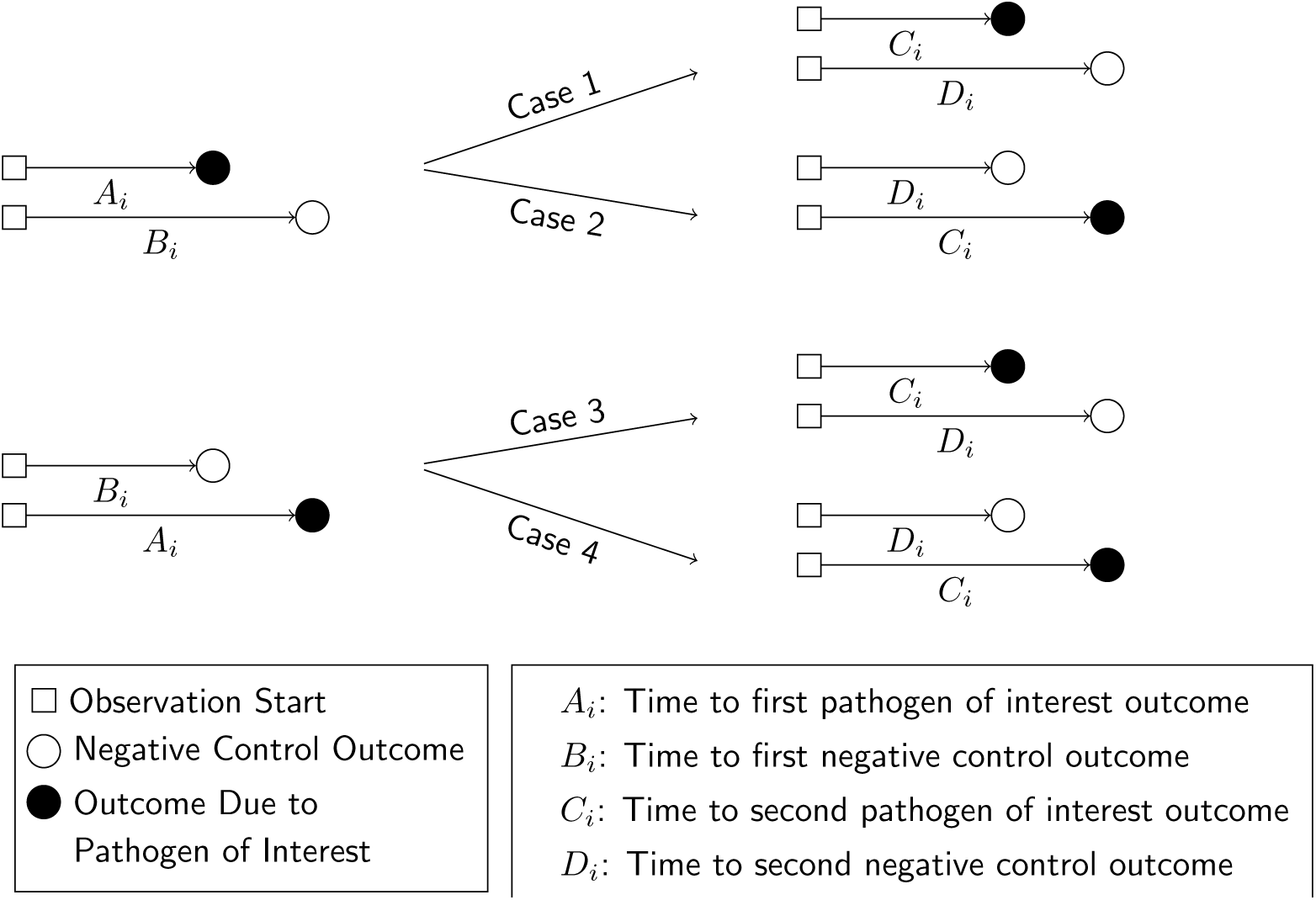
Schematic presentation of potential outcomes. We illustrate potential outcomes in terms of the sequence of events *A*_*i*_*-D*_*i*_ for a given individual. In cases 1 and 2, we observe *A*_*i*_ < *B*_*i*_ (truncating observation of a negative-control event while *X*_*i*_ = 0). In cases 3 and 4, we observe *B*_*i*_ < *A*_*i*_, with the negative-control outcome preceding infection with the pathogen of interest. We illustrate the corresponding potential outcomes for *C*_*i*_ and *D*_*i*_, when *X*_*i*_ = 1, in the right-hand side of the figure.

### Event time distributions

Define the hazard rate at which an individual *i* experiences the outcome of interest as *λ*_*Pi*_, and define *θ* as the hazard ratio of incidence of this outcome given previous infection (**Table 1**). Assuming events occur independently any time during the follow up, conditioning *λ*_*Pi*_ and *θ*, event times are exponentially distributed. Thus, the cumulative probability of experiencing the outcome by time *t*, for a previously-uninfected individual, is 1 − exp (−*λ*_*Pi*_ *t*), while the cumulative probability of experiencing the outcome by time *t*, had the same individual counterfactually been previously infected, is 1 − exp (−*θλ*_*Pi*_ *t*).

Consider that data are collected from each individual for endpoints besides the primary outcome of interest. Among these, suppose a negative control outcome occurs at a rate *λ*_*Ni*_ for individual *i*. This rate should be unaffected by individuals’ prior exposure to the pathogen of interest, according to the definition of a negative control in this context.^35^ Under the same assumptions, the probability of experiencing the negative control outcome by time *t*, for individual *i*, is 1 − exp (−*λ*_*Ni*_ *t*).

### Estimating the effect of naturally-acquired immunity

For an individual with no history of previous infection, consider the outcome of interest and the negative-control outcome to be competing risks. The events *E*_*i*_-*H*_*i*_ may be defined to indicate the relative ordering of event times *A*_*i*_-*D*_*i*_ according to the contingency structure presented in **Table 3**. Specifically, take *E*_*i*_ = *A*_*i*_ ≤ *B*_*i*_ and *G*_*i*_ = *C*_*i*_ ≤ *D*_*i*_ to indicate the outcome of interest precedes the negative-control outcome during the periods with *X*_*i*_ = 0 and *X*_*i*_ = 1, respectively. Define *F*_*i*_ = *B*_*i*_ < *A*_*i*_ and *H*_*i*_ = *D*_*i*_ < *C*_*i*_ as complements, where the negative-control outcome precedes infection or disease with the pathogen of interest while *X*_*i*_ = 0 and *X*_*i*_ = 1, respectively.

**Table 3:**
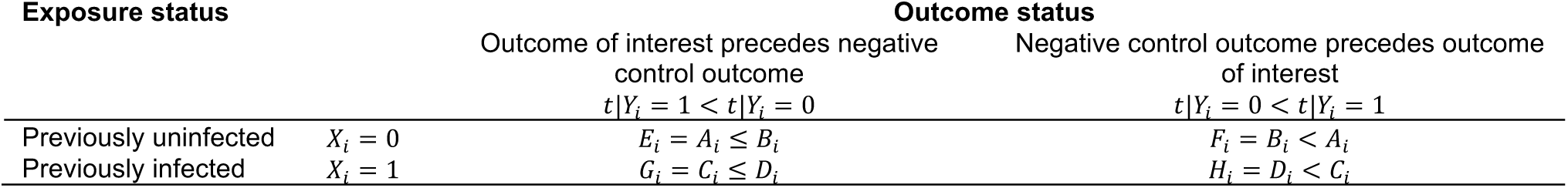
Contingency table for competing risks, given prior infection.

Under the scenario of exponentially-distributed event times (formulated in **Appendices A and B**), we have

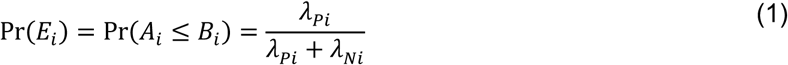

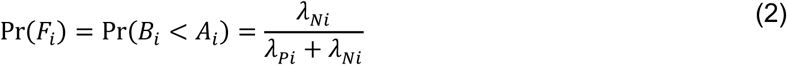

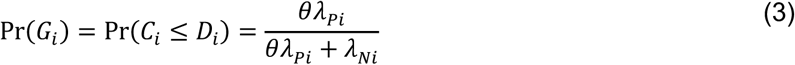

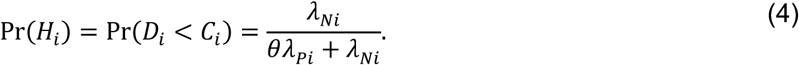

Consider the Mantel-Haenszel odds ratio^33,34^ constructed from the competing risks of *Y*_*i*_ = 1 and *Y*_*i*_ = 0, given *X*_*i*_, matching observations from each individual *i*:

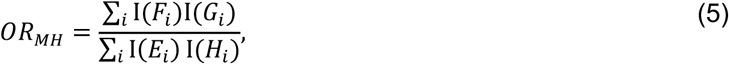

such that

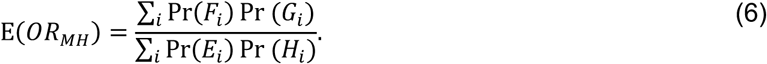

Using the above derivations of Pr(*E*_*i*_) through Pr(*H*_*i*_),

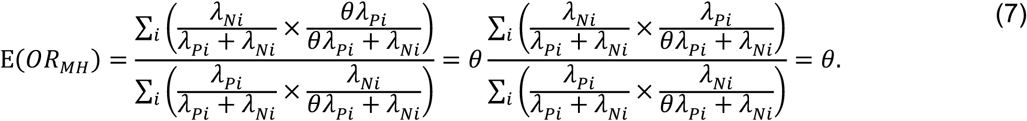

Thus, the ratio of the matched odds for the outcome of interest to precede a negative-control outcome, given an individual’s history of prior infection, provides an unbiased estimate of the effect of previous infection on time to recurrence of the outcome of interest.

### Further Considerations

At a design level, self-matched inference reduces or eliminates the potential for bias due to time-invariant factors that individually influence risk.^36^ However, complications arise when individuals’ risk of experiencing these endpoints differs substantially during the periods before and after individuals experience their first infection with the pathogen of interest.

To consider the implications of such time-varying confounding, define *β*_*Pi*_(0) and *β*_*Pi*_(1) as multipliers on the rate of infection with the pathogen of interest when *X*_*i*_ = 0 and *X*_*i*_ = 1, respectively, due to all factors other than infection-derived immunity against the pathogen of interest (**Table 2**). Similarly, define *β*_*Ni*_(0)and *β*_*Ni*_(1) as multipliers on the rate of the negative-control condition when *X*_*i*_ = 0 and *X*_*i*_ = 1, respectively, due to factors other than prior exposure to the pathogen of interest (**Table 4**). Here, *θβ*_*Pi*_ (1)/*β*_*Pi*_(0) and *β*_*Ni*_(1)/*β*_*Ni*_(0) are hazard ratios describing the relative incidence rate of the outcome of interest and the negative-control outcome, respectively, in the periods before and after infection with the pathogen of interest.

**Table 4:**
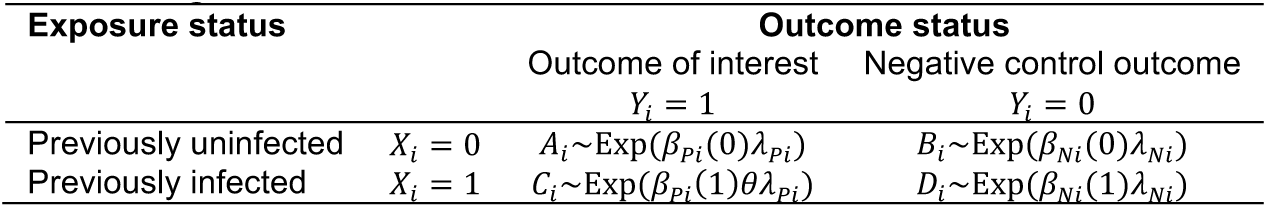
Contingency table for event time distributions, given prior infection, under the scenario of time-variant confounding.

| Exposure status |  | Outcome status |  |
| --- | --- | --- | --- |
|  |  | Outcome of interest | Negative control outcome |
| | | $Y_i = 1$ | $Y_i = 0$ |
| Previously uninfected | $X_i = 0$ | $A_i \sim \text{Exp}(\beta_{Pi}(0)\lambda_{Pi})$ | $B_i \sim \text{Exp}(\beta_{Ni}(0)\lambda_{Ni})$ |
| Previously infected | $X_i = 1$ | $C_i \sim \text{Exp}(\beta_{Pi}(1)\theta\lambda_{Pi})$ | $D_i \sim \text{Exp}(\beta_{Ni}(1)\lambda_{Ni})$ |

Here the matched odds ratio is

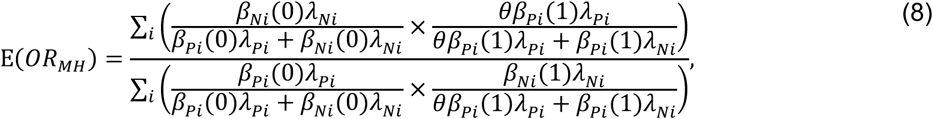

which reduces to *θ* only when *β*_*Pi*_(1)/*β*_*Pi*_(0) = *β*_*Ni*_(1)/*β*_*Ni*_(0) for all individuals. As we address in the **Discussion**, this circumstance motivates the selection of negative-control outcomes which resemble the outcome of interest in their association with time-varying confounders such as individuals’ age, health status, and sociodemographic exposures.

We may also consider a scenario where infection with the pathogen of interest alters individuals’ risk of the negative control endpoint, in addition their risk of reinfection or recurrent disease due to the pathogen of interest. Such a circumstance may arise if infection with the pathogen of interest causes individuals to modify risk behaviors that affect multiple outcomes, or confers broad (e.g. multi-pathogen) immunity.^37,38^

Defining *ρ* as the outcome-agnostic effect of infection with the pathogen of interest on future outcomes yields the contingency structure of **Table 5**.

**Table 5:** Two-by-two table for event time distributions, given prior infection, in the presence of pathogen-agnostic effects of previous infection.

| Exposure status |  | Outcome status |  |
| --- | --- | --- | --- |
|  |  | Outcome of interest | Negative control outcome |
| | | $Y_i = 1$ | $Y_i = 0$ |
| Previously uninfected | $X_i = 0$ | $A_i \sim \text{Exp}(\lambda_{Pi})$ | $B_i \sim \text{Exp}(\lambda_{Ni})$ |
| Previously infected | $X_i = 1$ | $C_i \sim \text{Exp}(\rho\theta\lambda_{Pi})$ | $D_i \sim \text{Exp}(\rho\lambda_{Ni})$ |

Here,

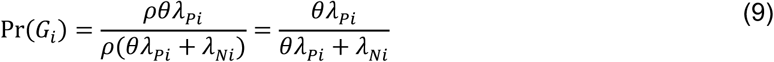

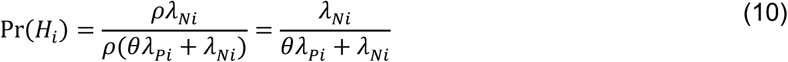

so the parameterization of 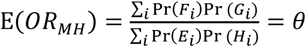 is unchanged. This circumstance is analogous to the more general case where *β*_*Pi*_(1)/*β*_*Pi*_(0) = *β*_*Ni*_(1)/*β*_*Ni*_(0)= *ρ*.

## COMPARISON TO COHORT DESIGN USING PROPORTIONAL HAZARDS ANALYSIS

### Simulation study

We conducted a simulation study across various underlying distributions of *λ*_*Pi*_ and *λ*_*Ni*_ to test for bias of point estimates under the proposed approach and under alternative methods often used in the analysis of cohort study data (without consideration of negative controls). As comparisons, we considered several proportional hazards models which could be applied to time-to-event data for recurrent observations of the outcome of interest. We considered four approaches to control for differences in hazard rates among individuals with differing exposure or susceptibility to the outcome of interest:

1. “Naïve” proportional hazards model without inclusion of additional terms to account for differences in event times among individuals. We define the hazard ratio estimated via fitting this model as 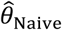.
2. Proportional hazards model accounting for variation in individual frailty via “random effects”. Fitting this model estimates the hazard ratio 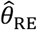 for the effect of previous infection, as well as 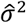 representing the estimated variance in (log) individual-specific event rates, assumed to represent independent draws from a Normal distribution with mean 0.^39,40^
3. Proportional hazards model including Gamma-distributed frailty terms.^13^ Fitting this model estimates the hazard ratio 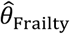 for the effect of previous infection, along with the parameters of the underlying Gamma distribution describing individual-specific frailties.
4. Proportional hazards model with “fixed effects” for individual subjects. Fitting this model estimates a hazard ratio 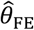 for the effect of previous infection and estimates subject-specific rates of infection (via individual-specific intercepts) which have no pre-specified distributional assumption.

We defined 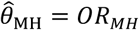 for the proposed analysis strategy of a self-matched, negative control design and considered various distributions for *λ*_*Pi*_:

1. Truncated Normal distribution (with a pre-specified lower bound at *a* = 0);
2. Truncated Cauchy distribution (with a pre-specified lower bound at *a* = 0);
3. Uniform distribution;
4. Gamma distribution;
5. Mixtures of Gamma distributions.

We considered multiple parameterizations of each of these distributions (**Table 6**), holding the mean rate (or location parameter of the Cauchy distribution) constant at one infection per year across all simulations to determine effects of inter-individual heterogeneity on estimates of *θ*. We illustrate the distributions in **Figure 3**. Considering cohorts of 500 individuals, we drew *λ*_*pi*_ values at random and sampled exponentially-distributed event times of first and second infections for each individual, truncating observations at five years. We repeated simulations 500 times for each *θ* ∈ {0.01, 0.02, … 0.99}, drawing *λ*_*Pi*_ values independently for each simulation. We used the simulated datasets to estimate 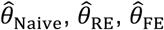, and 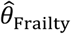, taking the average of estimates obtained across all 500 iterations to obtain a single point estimate for each parameterization.

**Table 6:**
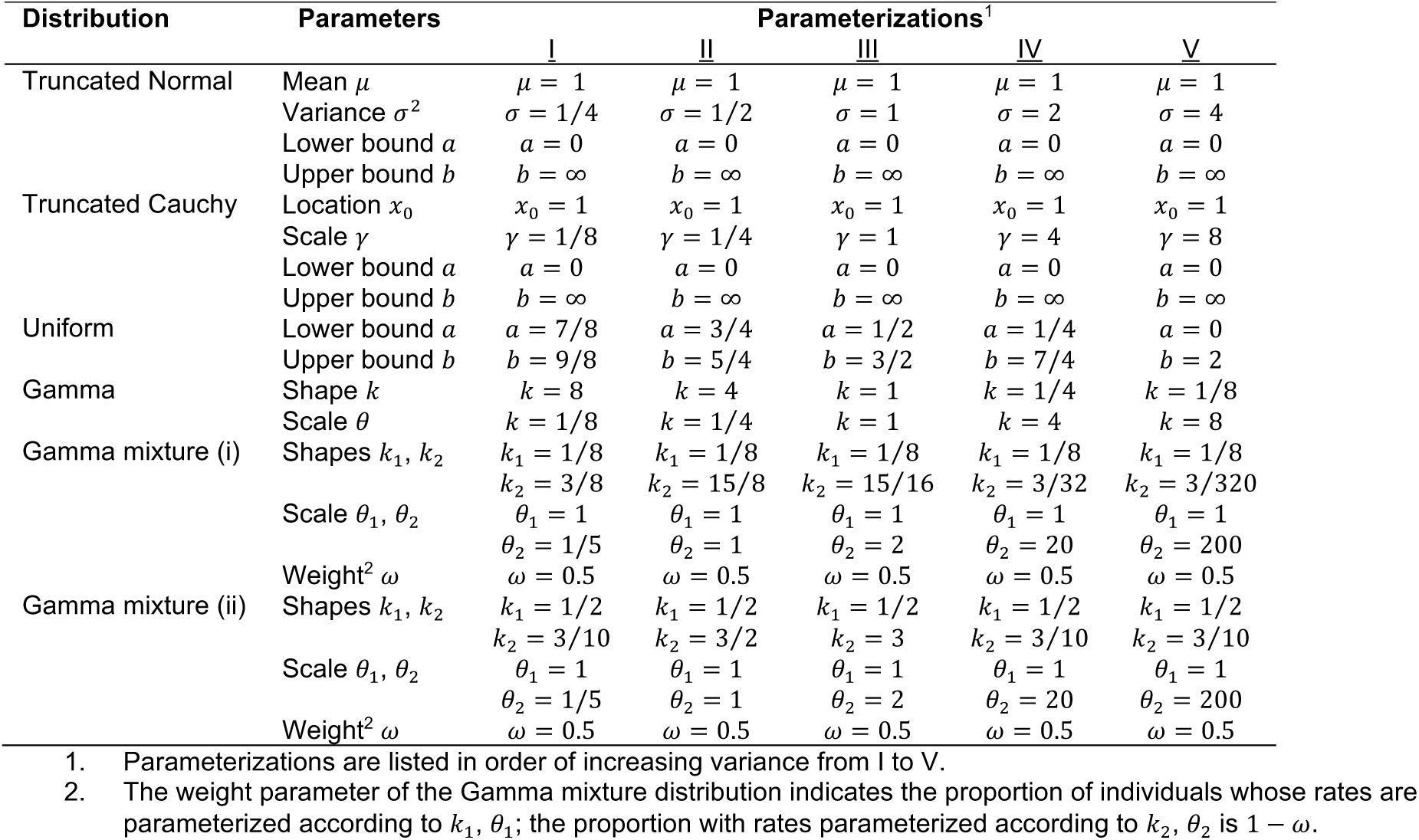
Event rate distributions applied to simulation study.

**Figure 3:**
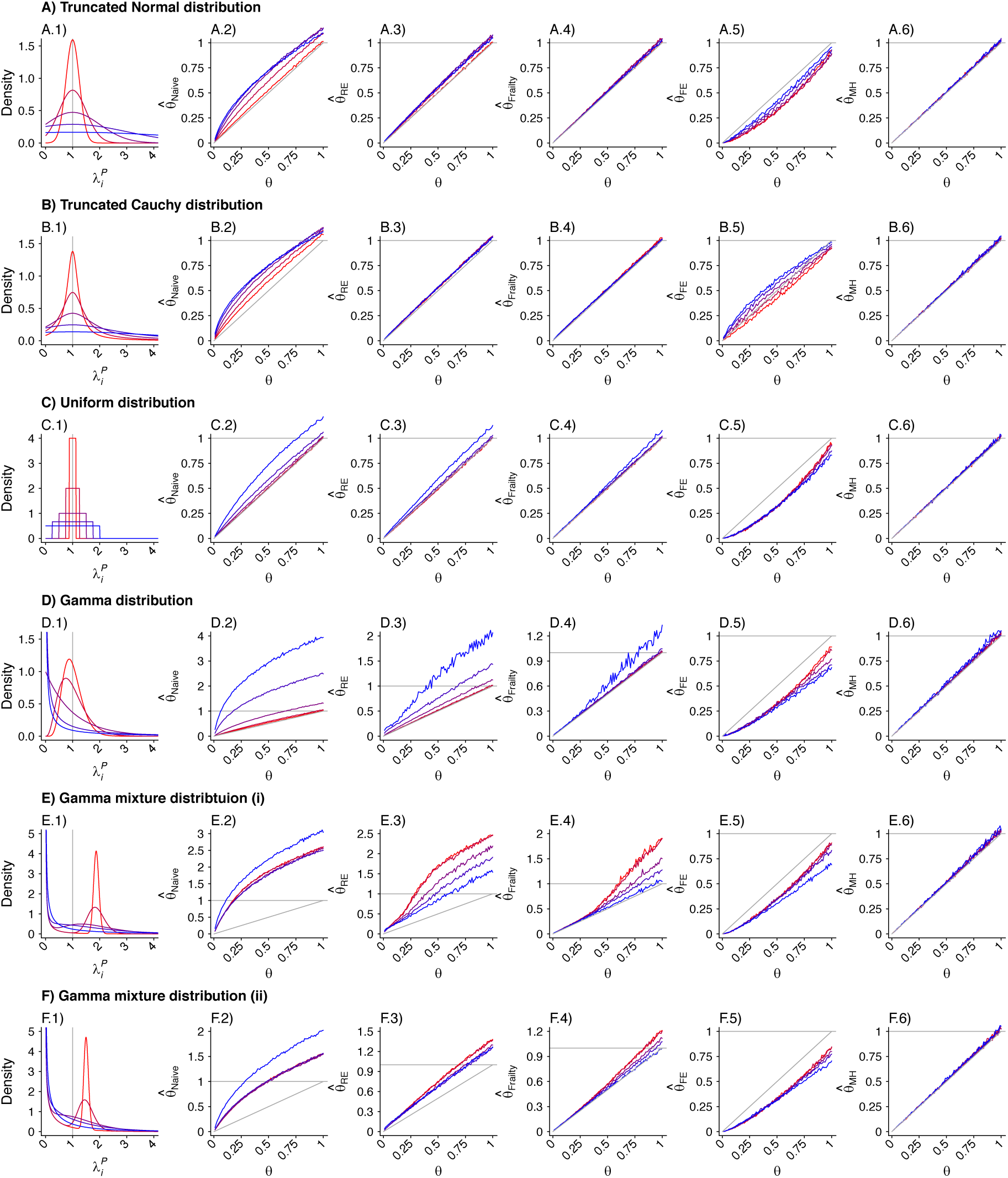
Simulated distributions and hazard ratio estimates under “naïve” inference approaches and under the proposed approach of self-matched inference with negative controls. Panels are organized to present, in each row, the assumed distribution (column 1), the estimate 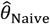 based on a Cox proportional hazards model without any correction for inter-individual heterogeneity (column 2), proportional hazards models employing various frailty frameworks (columns 3-5), and the estimate 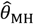. based on the proposed approach (column 6). One-to-one lines plotted in grey in columns 2-6 indicate where estimates would recover the true value, i.e. 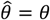. Horizontal grey lines plotted at 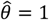 indicate where estimates exceed 1, indicating directionally-misspecified estimates of the causal effect of interest. Values are plotted on a red-to-blue color ramp corresponding to the parameterizations I-V, respectively, in order of least (I; red) to greatest (V; blue) variance as detailed in Table 4. **A**) Truncated Normal distribution; **B**) Truncated Cauchy distribution; **C**) Uniform distribution; **D**) Gamma distribution; **E**) Mixture of Gamma distributions (i) with means at 0.125 and 1.875; and **F**) Mixture of Gamma distributions (ii) with means at 0.5 and 1.5.

To compute 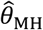, we drew rates (*λ*_*Ni*_) and event times for negative control observations from each subject, assuming event times were exponentially-distributed with respect to the underlying rates. To standardize comparisons of 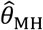 under differing distributions of *λ*_*Pi*_, we defined *λ*_*Ni*_ = 1 for all *i* under each simulation.

To investigate how the different modeling frameworks performed in capturing the distribution of individual-specific hazard rates, we saved estimates of individual-specific fixed effects, random effects, and frailties alongside estimates of 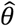. We fitted a single density kernel to the distribution of individual-specific estimates across 10 simulated cohorts for each true value of *θ* and underlying distribution of *λ*_*Pi*_.

## Results

We plot distributions and estimates under each approach in **Figure 3**. The naïve hazards ratio tended to overestimate *θ*, leading to under-estimation of the degree of protection (1 − *θ*). Bias was minimized as *θ* approached zero, consistent with a scenario of strong protective immunity. Values of 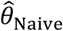 often exceeded 1 in scenarios where *θ* < 1; in practice, such an estimate would lead to inference that prior infection increases susceptibility to infection or disease due to the pathogen of interest, when in fact prior infection is protective. For all distributions considered, bias in 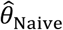 was greatest under parameterizations yielding the highest between-individual variance in *λ*_*Pi*_.

Alternative methods performed variably under the differing conditions (**Figure 3**). Lower degrees of bias were evident in 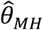 as compared to estimates generated under the other methods assessed. Gamma frailty models and random effects models tended to yield less-biased estimates of *θ* than 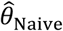. However, the same direction of bias (resulting in under-estimation of the reduction in susceptibility, or 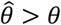) was evident with all three of these approaches. Bias was worst when *λ*_*pi*_ values were drawn from Gamma or Gamma mixture distributions, and tended to increase under distributions with greater variance in *λ*_*Pi*_, or greater irregularity in the case of Gamma mixture distributions. In contrast, fixed-effects models estimating multipliers on hazard rates for each individual tended to under-estimate *θ* under most distributions of *λ*_*Pi*_, although both 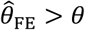 and 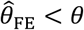 were apparent in simulations using the truncated Cauchy distribution for *λ*_*Pi*_. For the truncated Normal distribution, bias in 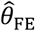 decreased with greater variance in *λ*_*Pi*_, whereas for the Uniform, Gamma, and Gamma mixture distributions, bias increased with greater variance in *λ*_*Pi*_.

Biased estimation of *θ* occurred in connection with a failure to accurately recover the underlying individual-specific frailty distributions. For each modeling approach, the extent of this misspecification in individual frailties varied over values of *θ* and distributions of *λ*_*Pi*_ (**Supplemental Digital Content, Figures S1-S3**).

## SAMPLE SIZE CONSIDERATIONS

### Simulation study

To inform applications of the proposed method, we next assessed statistical power under differing conditions. A test statistic (*ξ*_*MH*_) has previously been identified for *OR*_*MH*_ under the null hypothesis of no difference in risk given exposure.^41^ For the contingency structure (**Table 3**) formulated from the terms *E*_*i*_-*H*_*i*_ (by which we define *OR*_*MH*_), this statistic can be written generally as

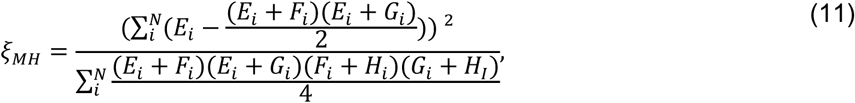

which can then be simplified according to *E*_*i*_ + *F*_*i*_ = 1, *G*_*i*_ + *H*_*i*_ = 1, and *F*_*i*_ + *H*_*i*_ = 2 − *E*_*i*_ − *G*_*i*_. Thus,

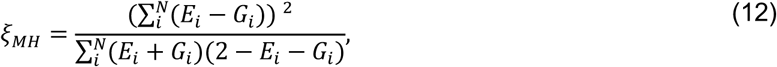

which is expected to follow a *χ*^6^ distribution with one degree of freedom under the null hypothesis. We calculated values of *ξ*_*MH*_ obtained for cohorts of varying sizes under differing parameterizations of *θ, λ*_*Pi*_, and *λ*_*Ni*_. For values of *θ* ∈ {0.1, 0.2, …, 0.9}, we sampled individual event times *A*_*i*_-*D*_*i*_ for a population of 100,000 individuals whom we subsequently partitioned (without replacement) into 2000 hypothetical study cohorts each of size *N=*25, 50, 75, 100, 150, 200, 250, 300, 350, 400, 450, 500, 600, 700, 800, and 1000. For these analyses, we considered *λ*_*Pi*_ values drawn from truncated Normal, Gamma, and Gamma mixture distributions, under the parameterizations of each of these distributions with greatest and least variance listed in **Table 6**. We determined statistical power via the proportion of simulated cohorts for which the upper bound of a 95% confidence interval around *OR*_*MH*_ would be expected to correctly exclude the null value, i.e. Pr 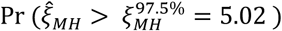).

To assess how correlation between *λ*_*Pi*_ and *λ*_*Ni*_ could affect the statistical power of estimates, we conducted simulations under two sets of assumptions. Under the first, we considered *λ*_*Ni*_ = 1 for all *i* (equal to the expected value of *λ*_*Pi*_ under all parameterizations), so that *λ*_*Ni*_ ⊥ *λ*_*Pi*_; under the second, we defined *λ*_*Ni*_ = *λ*_*Pi*_, under the assumption that individuals with greater risk of the outcome of interest would also experience higher incidence of the negative control condition. These conditions bound power estimates, corresponding to assuming no correlation and perfect correlation between *λ*_*Ni*_ and *λ*_*Pi*_, respectively.

## Results

We present results of the power analyses in **Figure 4**. Analyses with as few as 50 subjects had roughly 80% power or greater to estimate *θ* = 0.1 (corresponding to 90% protection) under all conditions explored; analyses with 500 subjects had 80% power or greater to estimate *θ* ≤ 0.5 (corresponding to 50% protection or greater) under all conditions. No scenarios revealed 80% or greater power for estimation of *θ* ≥ 0.8 (corresponding to less than 20% protection), even with 1000 subjects; statistical power for estimation of *θ* = 0.9 was 10% or lower under nearly all conditions explored.

**Figure 4:**
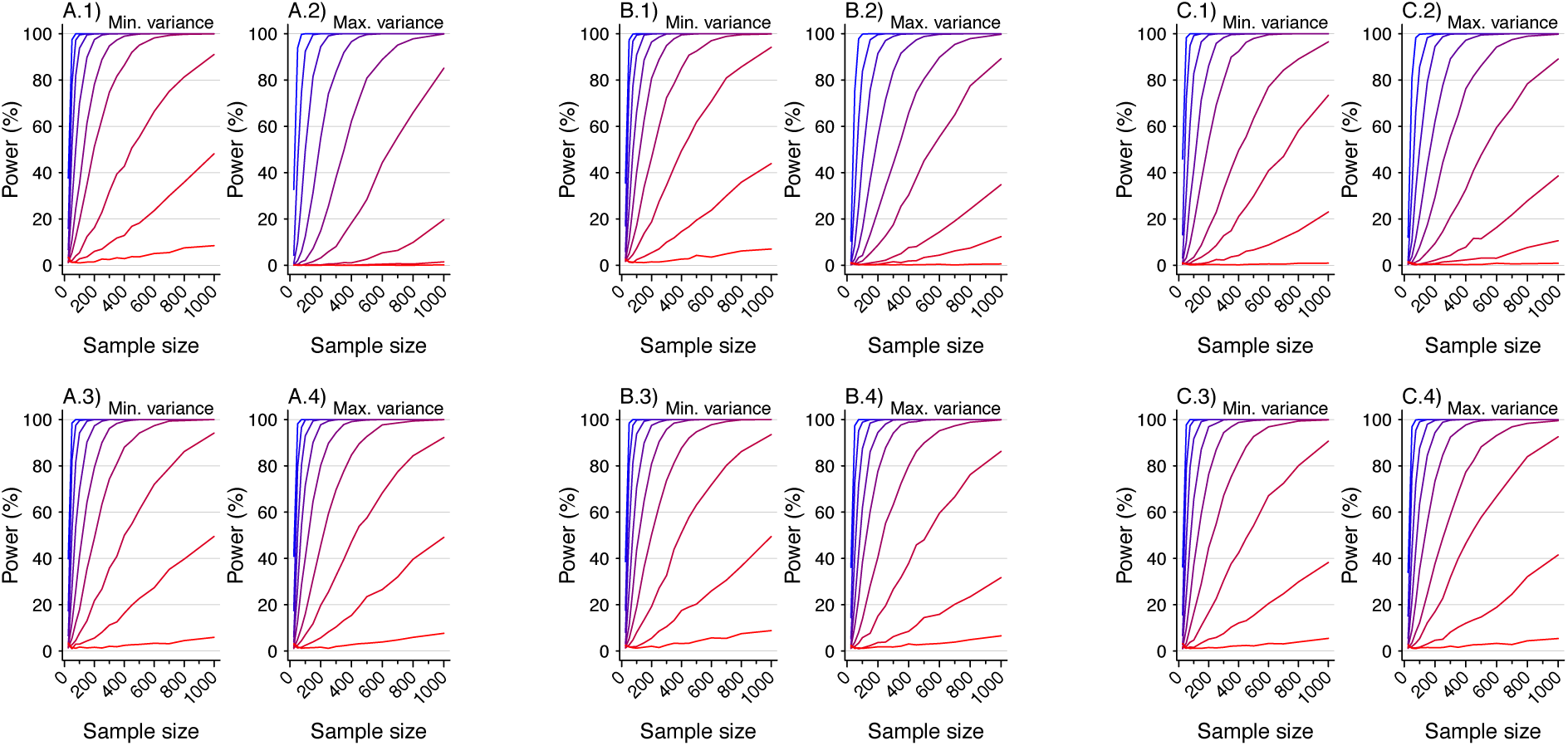
Statistical power for simulated analyses using the proposed approach of self-matched inference via negative controls. Each panel presents the statistical power for rejecting the null hypothesis with two-sided *p*<0.05 under varying conditions. Lines plotted in red to blue correspond to decreasing values of *θ*: 0.9 (red), 0.8, 0.7, …, 0.1 (blue), corresponding to increasing protection from 10% to 90%. Plots are presented in groups of 4 panels, each corresponding to analyses with values drawn from the following distributions: **A**) Truncated Normal distribution; **B**) Gamma distribution; **C**) Mixture of Gamma distributions with means at 0.125 and 1.875 (as detailed in **Table 4**). Panels in the top row (*A*.*1, A*.*2, B*.*1, B*.*2, C*.*1, C*.*2*) represent analyses in which no correlation is assumed between rates of the outcome of interest and negative control outcome (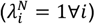). Panels in the bottom row (*A*.*3, A*.*4, B*.*3, B*.*4, C*.*3, C*.*4*) represent analyses in which the correlation between rates of the outcome of interest and the negative control outcome are is maximized (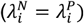). Within each grouping, panels on the left-hand side (*A*.*1, A*.*3, B*.*1, B*.*3, C*.*1, C*.*3*) correspond to distributions with the least variance in individual rates of the outcome of interest (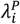; i.e., parameterization I in **Table 4**). Panels on the right-hand side within each grouping (*A*.*2, A*.*4, B*.*2, B*.*4, C*.*2, C*.*4*) correspond to distributions with the greatest variance in individual rates of the outcome of interest (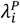; i.e., parameterization V in **Table 6**).

For simulations with *λ*_*Ni*_ ⊥ *λ*_*Pi*_, statistical power was weaker under parameterizations resulting in greater variance in *λ*_*Pi*_. In contrast, for simulations with *λ*_*Ni*_ = *λ*_*Pi*_, differences in statistical power were less strongly apparent with increasing variance in *λ*_*Pi*_. Taken together, these findings suggest statistical power is maximized when negative control endpoints are chosen which tend to occur more commonly among individuals who are at greatest risk of the outcome of interest.

## APPLICATION TO ROTAVIRUS BIRTH COHORT DATA

Last, we applied the proposed method to real-world data collected in two birth-cohort studies of rotavirus infection and disease among 200 children in Mexico City, Mexico and 373 children in Vellore, India. These datasets have been described extensively in primary study publications^4,5^ and subsequent re-analyses.^15,42^ Similar designs were employed for the studies. Briefly, pregnant mothers were enrolled prior to childbirth, and children were followed from birth to ages 2 years (in Mexico City) and 3 years (in Vellore). Investigators aimed to identify all rotavirus infections through routine testing of asymptomatic stool specimens (collected by field workers at regular home visits) for rotavirus, and by monitoring children for anti-rotavirus seroconversion over serial blood draws at scheduled intervals. Active surveillance was undertaken for all cases of gastroenteritis among children to characterize symptoms and test diarrheal stool specimens for rotavirus.

Initial analyses of the datasets led to differing conclusions about the strength of protection against rotavirus gastroenteritis (RVGE). Children in Mexico City were estimated to have experienced 77% (95% confidence interval: 60-88%), 83% (64-92%), and 92% (44-99%) lower rates of RVGE following one, two, and three previous infections, respectively, as compared to zero infections.^4^ In contrast, children in Vellore, where the rate of rotavirus acquisition was higher, were estimated to have experienced 43% (24-56%), 71% (59-80%), and 81% (69-88%) lower rates of RVGE after one, two, and three previous infections, as compared to zero infections.^5^ Subsequent analyses of the datasets revealed substantial variation in rates of rotavirus infection and risk of RVGE among individual children, as well as a potential for confounding due to declining risk of RVGE when infections were acquired at older ages, irrespective of previous infection.^42^ In contrast, model-based analyses accounting for the independent effects of age and previous infection on children’s susceptibility to RVGE estimated that children experienced 33% (23-41%), 50% (42-57%), and 64% (55-70%) lower rates of RVGE after one, two, and three previous infections, respectively, as compared to zero infections.^15^

We used the proposed self-matched negative control design to re-estimate naturally-acquired protection against RVGE in the cohort datasets. Here, RVGE episodes (acute, new-onset diarrhea with rotavirus detected in the stool) are the outcome of interest and acute, new-onset diarrhea episodes without rotavirus detection as the negative control. We compared the times of RVGE and rotavirus-negative diarrhea episodes from each child beginning from birth, and thereafter following detection of the first, second, and third rotavirus infection (generating confidence intervals via resampling of individual children). This yielded estimates of 27% (–1-48%), 50% (13-73%), and 48% (0-77%) lower rates of RVGE following one, two, and three previous infections, as compared to zero infections (**Figure 5**). Notwithstanding lower statistical power for the proposed method, these estimates are in agreement with previous findings^15^ suggesting lower strength of naturally-acquired protection than what was estimated in initial analyses of the birth cohort studies.^4,5^

**Figure 5:**
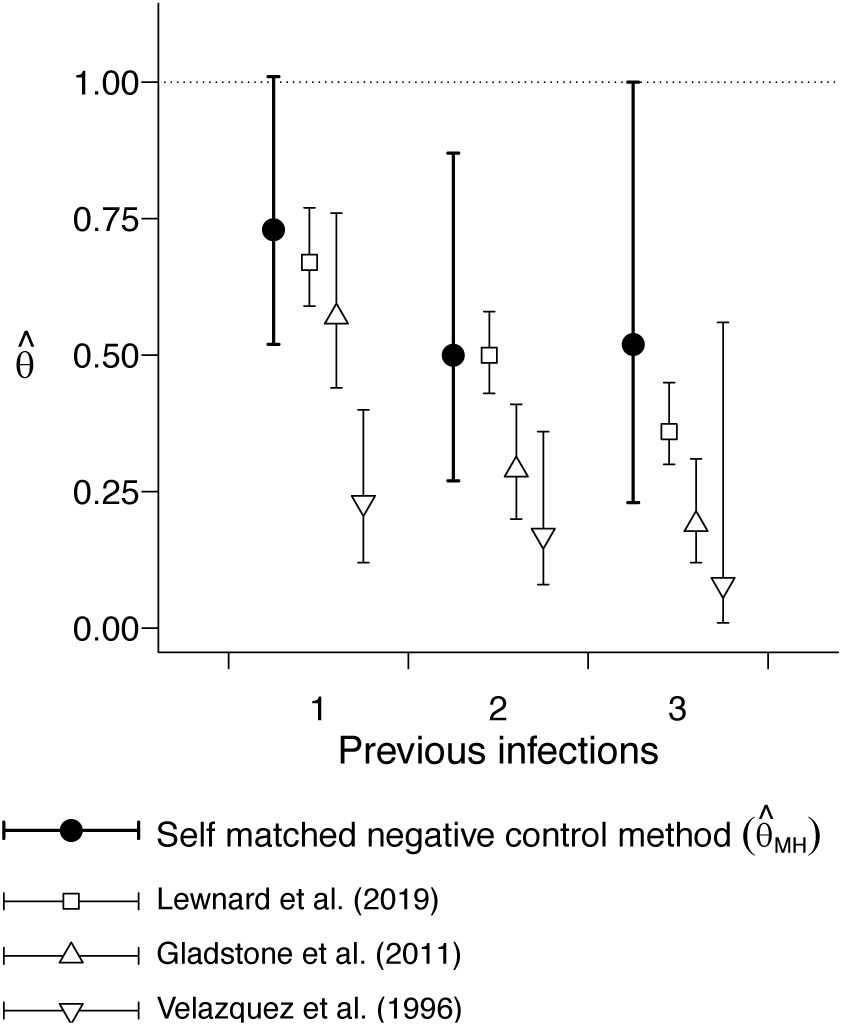
Estimated protection against rotavirus gastroenteritis associated with previous infection. We plot point estimates and 95% confidence intervals (lines) for estimates of the hazard ratio of rotavirus gastroenteritis associated with having previously experienced one, two, and three previous infections, versus zero previous infections, estimated via re-analysis of the Mexico City and Vellore rotavirus birth cohort studies.^4,5^ Analyses include rotavirus-negative diarrhea occurrences as a negative control endpoint.

## DISCUSSION

We propose a novel self-matched negative control method for estimating the hazard ratio of time to infection or disease due to a pathogen of interest, given previous infection. Analytically and via simulation, we show this method recovers unbiased estimates under a range of conditions, including when individual incidence rates of the outcome of interest are drawn from highly irregular or skewed distributions. We find these irregular or skewed distributions may lead to bias under proportional hazards models with commonly-used frailty estimation frameworks. Desirably, the proposed approach requires no parametric assumptions other than event-times being exponentially distributed with respect to their underlying, individual-specific rates of occurrence. Beyond infectious disease natural history studies, this approach may have value for assessing the effects of other exposures on recurrent event times.

Our findings provide several practical insights for real-world longitudinal cohort studies. Collecting data on multiple endpoints affords the opportunity to leverage negative-control observations to support causal inference. For studies applying the proposed approach, negative-control endpoints affected by the same risk factors or exposures as the outcome of interest are desirable both to reduce potential risks of confounding due to time-varying factors, and to maximize statistical power based on the correlation between event rates for the outcome of interest and negative-control outcome, *λ*_*Pi*_ and *λ*_*Ni*_. “Test-negative” control conditions which resemble the outcome of interest, but are not attributable to the same pathogen,^43,44^ may provide a compelling choice, particularly if their occurrence is predicted by similar risk factors. For instance, shared risk factors are well-documented for rotavirus-positive and rotavirus-negative diarrhea.^15,31^ Considering respiratory illness, multiple etiologic viruses may share similar seasonal transmission patterns,^45^ routes of transmission via high-risk contact,^46^ and associations of disease progression or severity with host comorbidities.^47^ For sexually-transmitted infections, particular risk behaviors^48^ differing among individuals or over time could alter risk of any infection, rather than infection with the pathogen of interest alone.^25^ In the context of real-world cohort studies, test-negative control conditions which are clinically similar to the outcome of interest would likely result in a study visit or other recorded interaction with similar probability. This further supports consideration of inference methods making use of test-positive and test-negative occurrences of a particular clinical syndrome.

In summary, self-matched inference via negative controls may provide a flexible strategy to circumvent bias introduced by variation in individual frailty for analyses of naturally-acquired immunity. Applications to other exposures affecting the distribution of recurrent event times merit consideration, given the possible limitations we identify in existing analysis frameworks.

## Data Availability

Code for replication of the analyses is available at github.com/gnorthrup/SelfMatchedNegativeControl.

## APPENDIX A COMPETING RISKS

For two competing, independent event times *τ*_*j*_ and *τ*_*l*_ occurring at rates *r*_*j*_ and *r*_*k*_, the probability for *τ*_*j*_ to precede *τ*_*k*_ is

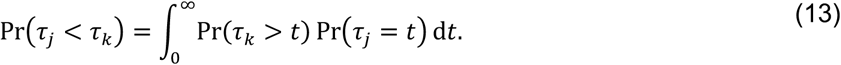

Under the assumption of exponentially-distributed event times,

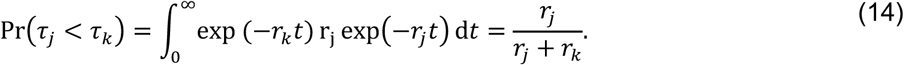

## APPENDIX B TRUNCATION OF OBSERVATIONS

The derivation above considers the indefinite integral

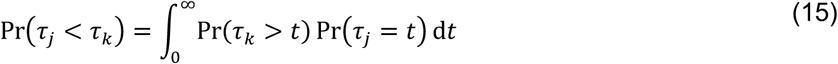

We obtain the same results when considering bounded observations truncated at time *δ*, more in line with the conduct of real-world studies:

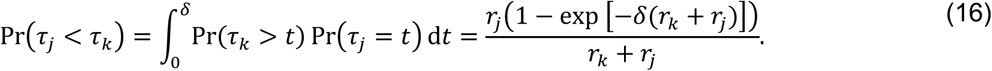

Thus,

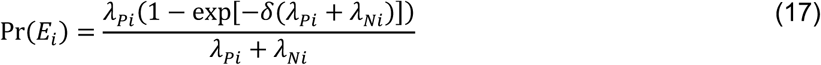

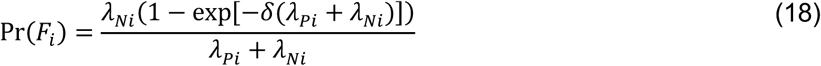

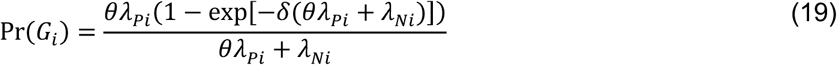

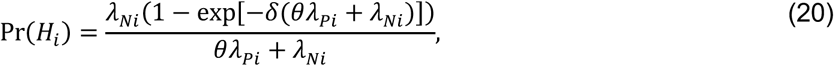

with the additional terms cancelling out in the matched odds ratio formulation:

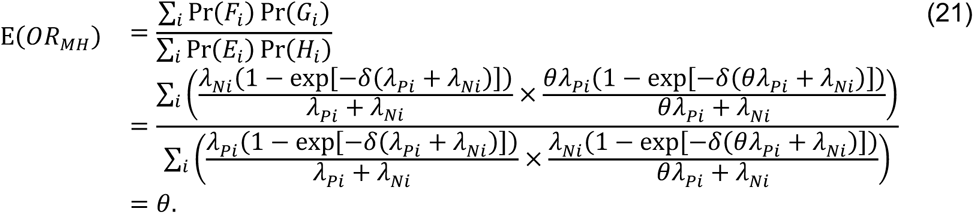

---

**FIGURE S1:**
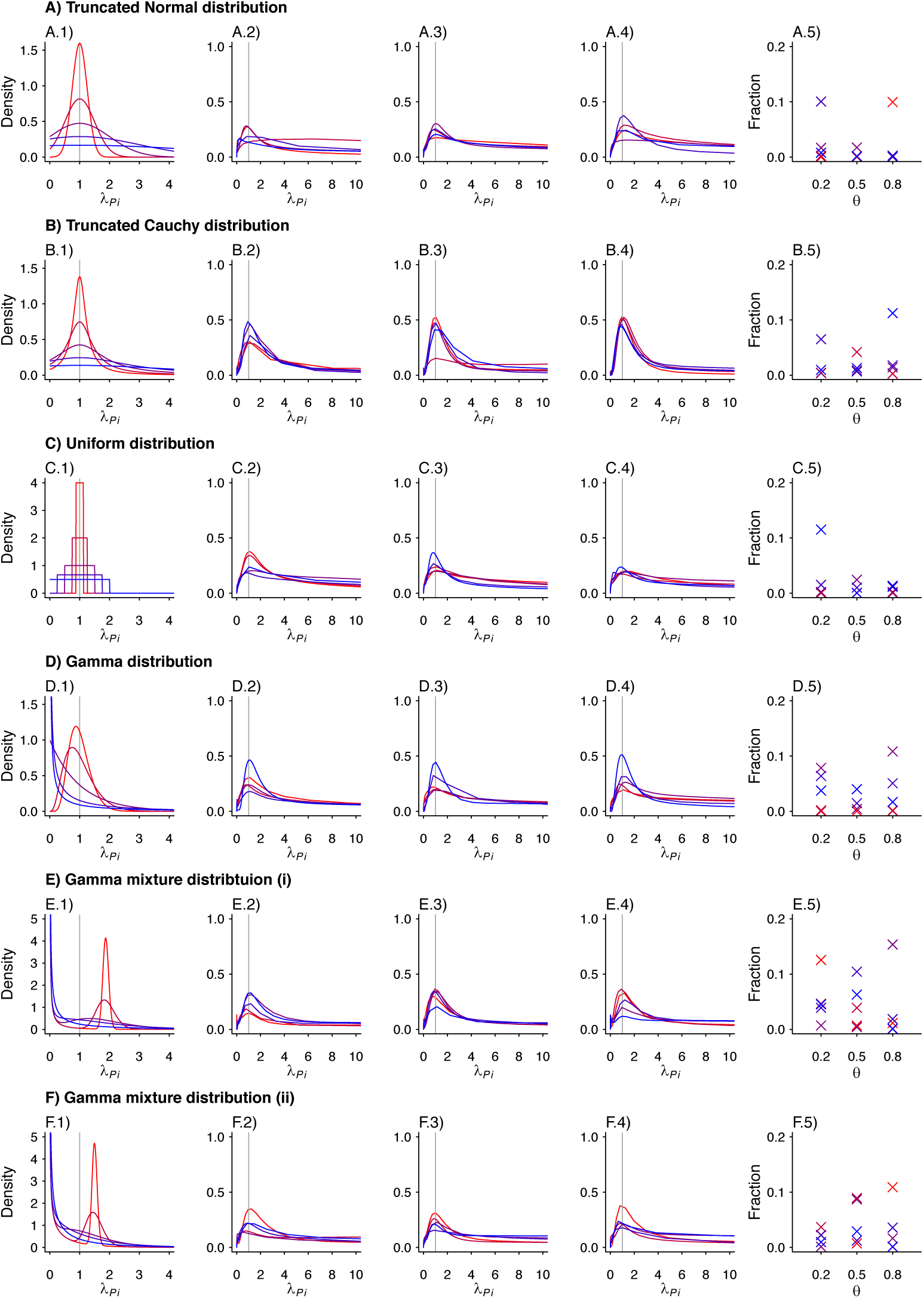
Estimated Density Kernels For Fixed Effects Model. We plot the estimated density kernels for the individual effects estimated by the fixed effects model for each individual in a simulated study. In each row, column 1 is a reproduction of the densities used to produce the individual *λ*_*Pi*_. Columns 2-4 are the estimated density kernels for *θ* = 0.2, *θ* = 0.5, *θ* = 0.8, respectively. Column 5 shows the proportion of estimates which were over 1000 for each set of parameters. Values are plotted on a red-to-blue color ramp corresponding to the parameterizations I-V, respectively, in order of least (I; red) to greatest (V; blue) variance as detailed in Table 4. **A**) Truncated Normal distribution; **B**) Truncated Cauchy distribution; **C**) Uniform distribution; **D**) Gamma distribution; **E**) Mixture of Gamma distributions (i) with means at 0.125 and 1.875; and **F**) Mixture of Gamma distributions (ii) with means at 0.5 and 1.5.

**FIGURE S2:**
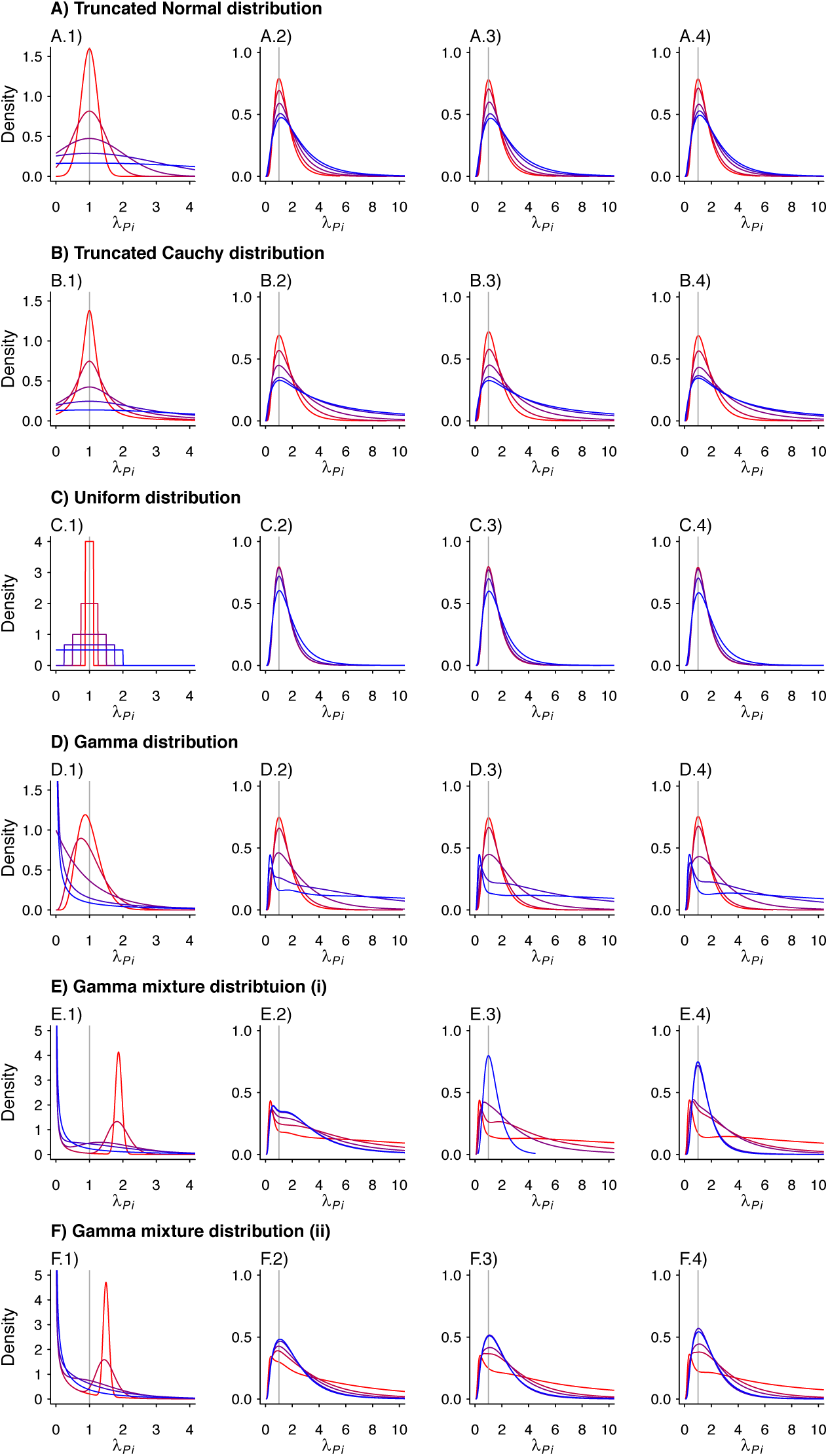
Estimated Density Kernels For Random Effects Model. We plot the estimated density kernels for the individual effects estimated by the random effects model for each individual in a simulated study. In each row, column 1 is a reproduction of the densities used to produce the individual *λ*_*Pi*_. Columns 2-4 are the estimated density kernels for *θ* = 0.2, *θ* = 0.5, *θ* = 0.8, respectively. Values are plotted on a red-to-blue color ramp corresponding to the parameterizations I-V, respectively, in order of least (I; red) to greatest (V; blue) variance as detailed in Table 4. **A**) Truncated Normal distribution; **B**) Truncated Cauchy distribution; **C**) Uniform distribution; **D**) Gamma distribution; **E**) Mixture of Gamma distributions (i) with means at 0.125 and 1.875; and **F**) Mixture of Gamma distributions (ii) with means at 0.5 and 1.5.

**FIGURE S3:**
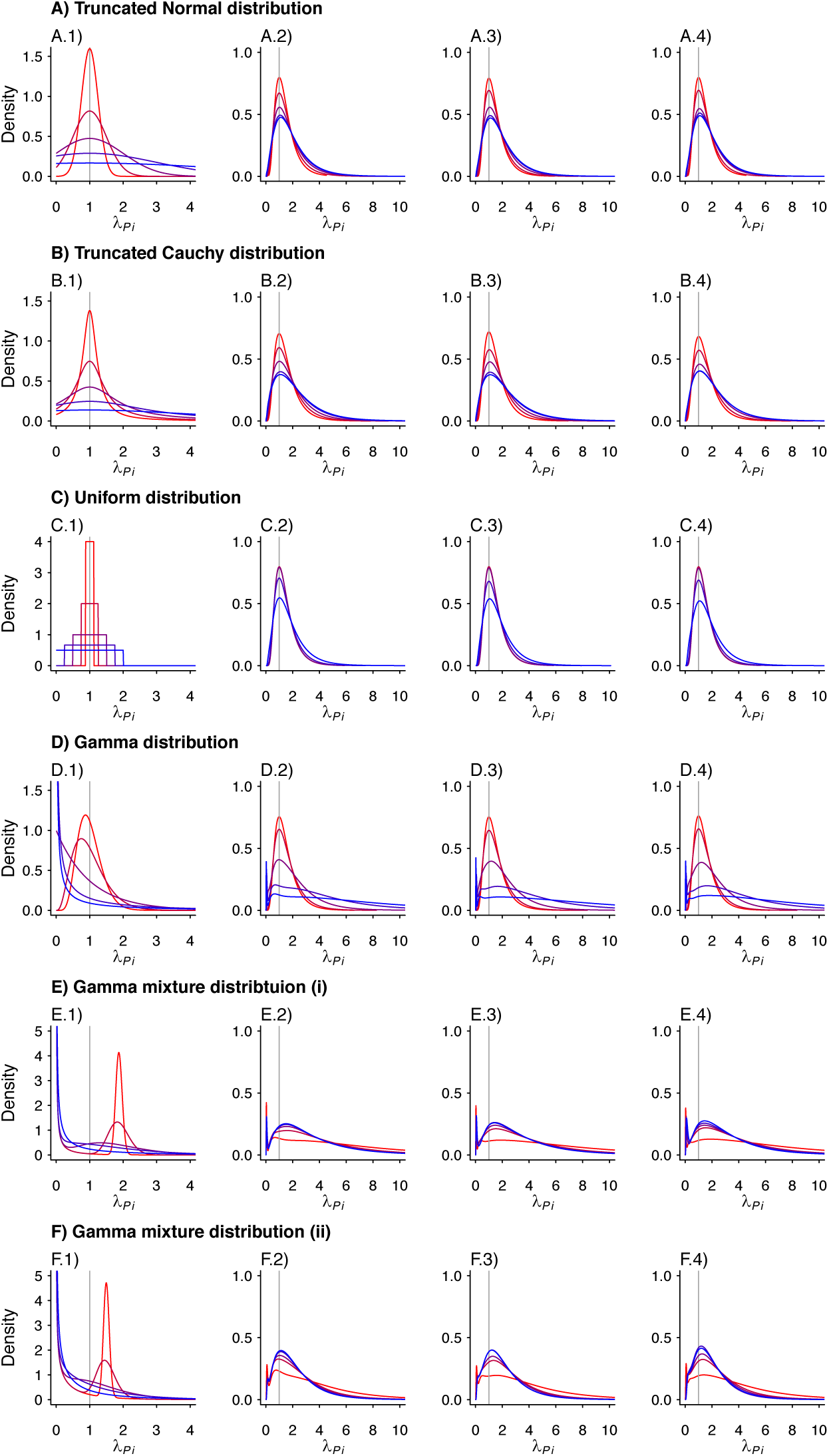
Estimated Density Kernels For Gamma Frailty Model. We plot the estimated density kernels for the individual effects estimated by the gamma frailty model for each individual in a simulated study. In each row, column 1 is a reproduction of the densities used to produce the individual *λ*_*Pi*_. Columns 2-4 are the estimated density kernels for *θ* = 0.2, *θ* = 0.5, *θ* = 0.8, respectively. Values are plotted on a red-to-blue color ramp corresponding to the parameterizations I-V, respectively, in order of least (I; red) to greatest (V; blue) variance as detailed in Table 4. **A**) Truncated Normal distribution; **B**) Truncated Cauchy distribution; **C**) Uniform distribution; **D**) Gamma distribution; **E**) Mixture of Gamma distributions (i) with means at 0.125 and 1.875; and **F**) Mixture of Gamma distributions (ii) with means at 0.5 and 1.5.

